# Presence of *Dolosigranulum pigrum* in the nasopharynx and its relationship with respiratory health status in paediatric population

**DOI:** 10.64898/2026.03.13.26347700

**Authors:** M Cisneros, D Henares, A Lluansi, P Brotons, C Launes, D Penela-Sanchez, G Gonzalez-Comino, A Perez-Arguello, MF de Sevilla, A Mira, M Blanco-Fuertes, C Muñoz-Almagro

## Abstract

**Background:** Respiratory tract infections range from asymptomatic colonisation to an invasive disease. Recent studies suggest that nasopharyngeal microbiota may influence this variability. Emerging evidence points to *Dolosigranulum pigrum,* a nasopharyngeal commensal, as a potentially protective bacterium. This study aimed to identify variables associated with the presence of *D. pigrum* in the nasopharynx of children with varying respiratory health statuses.

**Methods:** Nasopharyngeal aspirates were collected from children <18 years who were asymptomatic (n=65), had banal viral infection (n=48), or Invasive Pneumococcal Disease (IPD) (n=27). The presence of *D. pigrum* was defined as >0.1% of total sequences obtained by 16S rRNA gene sequencing. Variables included sex, breastfeeding, delivery mode, *S. pneumoniae* carriage, respiratory viruses and clinical features.

**Results:** Among 140 children (73 males, 67 females), *D. pigrum* was detected in 79 (56.4%): 44/65 in the healthy group; 26/48 of viral and 9/27 IPD cases. Multivariate analysis revealed significant associations with health status and sex. Healthy children were more likely to carry *D. pigrum* than IPD cases (44/79 vs. 26/79; p= 0.028). Males were more frequently *D. pigrum* carriers than females (48/79 *vs.* 31/79; p= 0.033).

**Conclusion:** *D. Pigrum* was associated with respiratory health, being more prevalent in healthy children, and showed potential sex-related differences.

## INTRODUCTION

Respiratory tract infections (RTIs) remain a major public health challenge, with lower respiratory tract infections (LRTIs) being the leading cause of death worldwide, especially in children under five years [1]. These infections are commonly caused by pathobionts such as *Streptococcus pneumoniae, Haemophilus influenzae, Staphylococcus aureus,* or by respiratory viruses. Pathobionts possess a dual potential role: they can asymptomatically colonise the nasopharynx, but under certain conditions, they may transition to causing local diseases or invade normally sterile anatomical sites, leading to invasive disease [2]. *S. pneumoniae* is one major respiratory pathogen due to the high burden of invasive pneumococcal disease (IPD) [2,3].

Emerging evidence suggests that the human respiratory microbiota composition could be an important factor influencing the different behaviours of nasopharyngeal pathobionts [4]. The respiratory microbiota comprises diverse microbial communities that colonise the respiratory tract. These communities appear to play an important role in protecting against or increasing the risk of some infectious diseases. Nasopharyngeal microbiome dysbiosis, characterised by a decrease in microbial diversity and depletion of commensal bacteria that normally prevent pathogenic overgrowth, can lead to increased susceptibility to infection [5–7]. Specifically, certain studies have identified the *Corynebacterium* and *Dolosigranulum* species as significant beneficial members of the nasopharynx microbiota [8–14].

*Dolosigranulum pigrum* first described by Aguirre, et al. in 1993 [15] is a commensal bacterium from the upper respiratory tract commonly isolated from the nasal cavity and nasopharynx [15]. This gram-positive coccus is a lactic acid bacterium that is normally susceptible to beta-lactams [16]. Extensive evidence supports *D. pigrum* as a beneficial respiratory commensal. Microbiota studies have reported that respiratory health is strongly associated with the detection of the genus *Dolosigranulum* in the nasopharynx and the nasal cavity [17,18]. Conversely, absence or low abundance of *D. pigrum* has been linked to a wide variety of respiratory infections, including otitis, sinusitis, bronchitis, bronchiolitis, and pneumonia, as well as exacerbations of chronic respiratory diseases. In particular, our group has demonstrated this association extended beyond respiratory infections to invasive disease caused by *S. pneumoniae* [19,20].

Despite epidemiological evidence linking *D. pigrum* to respiratory health, the mechanisms remain poorly understood. It has been postulated an antagonism with the main respiratory pathogens. In vitro studies have demonstrated inhibition of pneumococcal growth by *D. pigrum*, though specific mechanisms have not been fully elucidated [14,21]. Additionally, *D. pigrum* appears to modulate respiratory innate immunity and enhance resistance to pathogens such as Respiratory Syncytial Virus (RSV) and S*. pneumoniae* [22–25]. Notably, nasal administration of *D. Pigrum* has been shown to decrease the number of *S. pneumoniae* and reduce its spread to the blood [23].

In light of the substantial evidence supporting the potential benefits and protective role of *D. Pigrum* in relation to respiratory health, it is essential to understand the factors that influence its presence in the nasopharynx. Therefore, the aim of the present study was to identify and analyse the epidemiological, clinical and microbiological variables that may be associated with the presence or absence of *D. pigrum* in the nasopharynx of children under the age of 18 years with different respiratory health statuses.

## MATERIAL AND METHODS

### Study design

This ancillary study used data from a previously case-control study at Sant Joan de Déu Barcelona Children’s Hospital (HSJD) from January 2014 to December 2018. The cohort included three paediatric groups according to health status: (i) healthy/asymptomatic outpatients; (ii) outpatients with microbiologically verified banal symptomatic viral RTIs; and (iii) inpatients with IPD. No statistically significant differences were observed among groups regarding age, sex, or seasonality [20].

The present analysis did not involve new participant recruitment or additional sample collection. Data for the current secondary analysis were accessed in 2025. The dataset was de-identified prior to analysis, and the authors did not have access to information that could identify individual participants.

The inclusion criteria were: (i) meeting the case/control definition previously detailed [20] (ii) having informed consent signed by the parents or legal guardians of participants; (iii) participants not belonging to a previously defined clinical risk group for developing IPD [26]; and (iv) no antibiotic exposure before sample collection (or ≤24 h for IPD cases) [27,28].

### Data collection

Diverse epidemiological, clinical, and microbiological variables were collected from all participants. Epidemiological variables included age, sex, ethnicity, delivery mode, maternal breastfeeding, pneumococcal vaccination status, and others. Clinical parameters were only applicable to IPD cases and included length of hospital stay, admission and length to the Paediatric Intensive Care Unit (PICU), and clinical manifestations. Microbiological parameters included pneumococcal colonisation status, pneumococcal nasopharyngeal load, invasive disease potential of nasopharyngeal pneumococcal serotypes, composition of the nasopharyngeal bacterial microbiota, and respiratory viral detection. All the variables collected were previously described [20].

### Sample collection

Nasopharyngeal aspirates (NPAs) were collected from all the participants included in the study. The procedure to collect these samples was previously published [20]. The samples were kept at -80°C until the laboratory analyses were conducted.

### Pneumococcal detection, quantification, and serotyping

A duplex real-time PCR targeting the *lytA* gene of *S. pneumoniae* and RNase P, a human control gene that identifies sample viability and inhibitors, was used in NPAs samples for detection of *S. pneumoniae*. Primers and probes were utilised according to the Centers for Disease Control and Prevention (CDC) guidelines [29].

### Respiratory virus detection

The multiplex real-time PCR Allplex II RV16 detection kit (Seegene) was used to detect the DNA/RNA of 15 of the most frequent human respiratory viruses on NPAs [30].

### Variable definition

In the current study, the main outcome was the presence or absence of *D. pigrum* in NPA, which was determined by bacterial 16S rRNA gene sequencing in the preceding main study [20]. The sequences assigned to *Dolosigranulum spp*. were analysed and compared to the total sequences to calculate the relative abundance of *D. pigrum. D. pigrum* was considered to be present in NPAs if it comprised >0.1% of the total sequences. The presence of *D. pigrum* was correlated with epidemiological, clinical, and microbiological variables previously collected. The serotypes 1, 3, 4, 5, 7F, 8, 9A, 9V, 12F, 14, 18C, 19A, and 33F were considered as high-invasiveness disease potential serotypes, as previously reported [20].

For subanalyses of relative abundance of *D. pigrum*, the healthy group was further stratified based on respiratory virus detection status into: (i) healthy virus-negative (asymptomatic without respiratory virus detection) and (ii) healthy virus-positive carriers (asymptomatic with respiratory virus detection).

### Statistical analyses

Statistical analyses were performed using the 4.4.2 version of R and RStudio software [31] using *car* [32], MASS [33], *emmeans* [34] and *lmtest* [35] packages. Shapiro-Wilk test was used to evaluate the normality of the data. Significance of associations between the presence or absence of *D. Pigrum* with categorical variables was tested by Chi-Square or Fisher’s test. Fisher’s test was used if ≥25% of cells had expected frequencies <5. Associations of *D. Pigrum* presence or absence with continuous variables were analysed using the Student t-test or the Mann-Whitney test for normally or not normally distributed data, respectively.

In order to assess potential relationships between the epidemiological, clinical, and microbiological variables and the presence of *D. pigrum*, a generalised linear model (GLM) with a binomial distribution and a logistic link function was used. An initial model was built up, including all the variables that showed an association with the presence of *D. pigrum* at a p-value ≤0.2 and did not have collinearity. A stepwise approach combining forward and backward selection was used to gradually add and eliminate variables to refine the optimal model. This strategy was based on the Akaike Information Criterion (AIC), which balances model fit and complexity [36]. To determine collinearity among the variables, the Variance Inflation Factor (VIF) was calculated, and the variables with a VIF >5 were considered collinear and excluded from the final model [37].

As a complementary quantitative analysis to assess the effect of abundance beyond presence/absence (qualitative analysis), the relative abundance of *D. pigrum* across respiratory health status groups was evaluated by the Kruskal-Wallis test, followed by the pairwise Wilcoxon test. This analysis included all the participants. Relative abundance values were log10-transformed prior to analysis.

Associations were measured by adjusted Odds Ratios (aORs) with corresponding 95% confidence intervals (CI). Estimated Marginal Means assessed the significance of the associations. A p-value <0.05 was established to determine statistical significance.

## RESULTS

### Characteristics of study population

A total of 140 participants were selected for the study, of which *D. pigrum* was detected in 79 (56.4%) (median age: 33.6 months [IQR: 2.0–41.5]) and absent in 61 (median age: 38.5 months [IQR: 0.1–48.0]). Detection rates were similar between children under two years of age (36/62, 58.1%) and older children (43/78, 55.1%). Regarding respiratory health status, *D. pigrum* was found in 44/67 (65.7%) in healthy children, 26/48 (54.2%) in viral cases and 9/27 (33.3%) in IPD cases *(Fig. 1A, Table 1).* The proportion of *D. pigrum* carriers differed significantly across healthy and IPD groups (p= 0.002).

**Fig 1.**
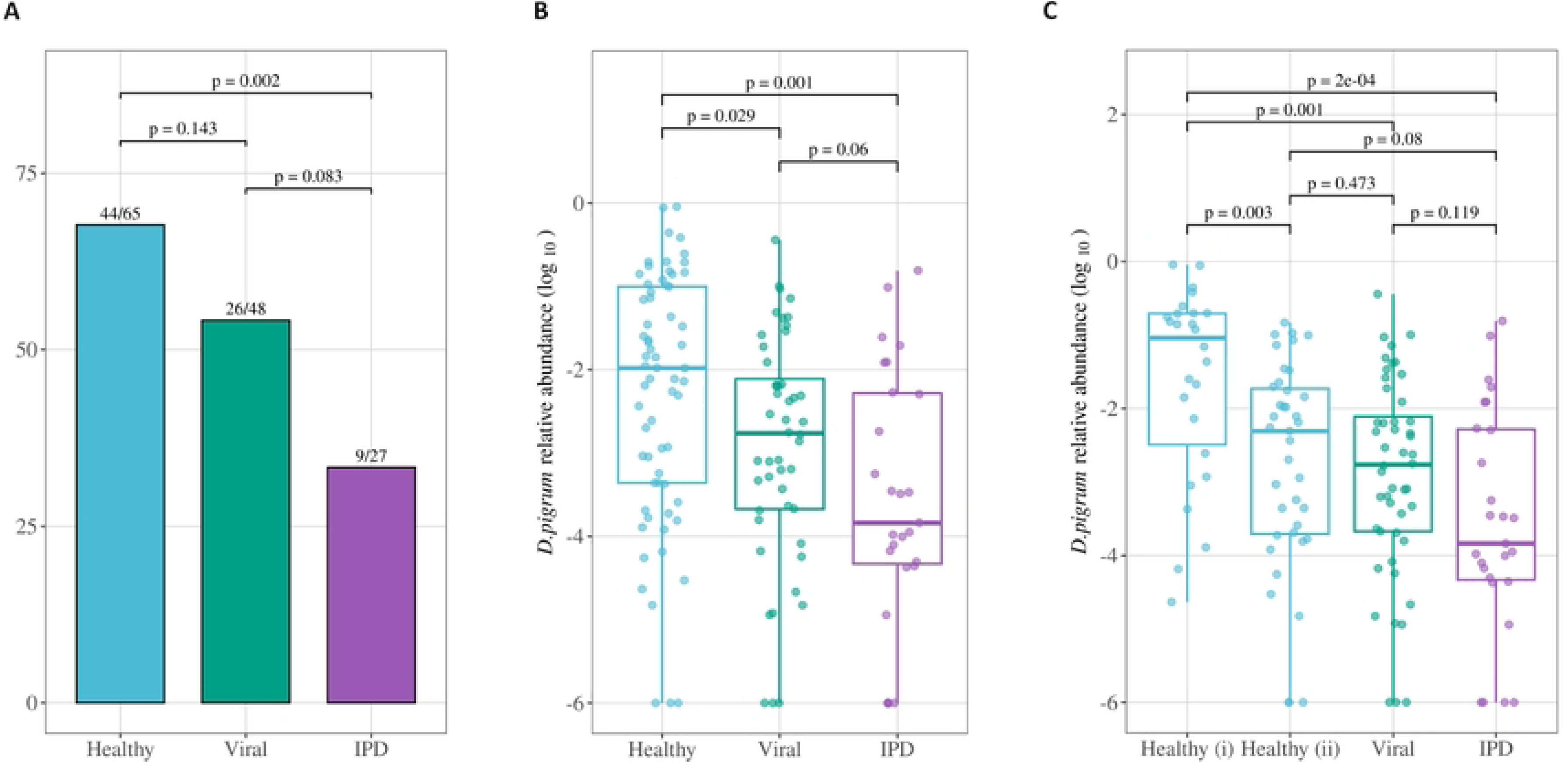
This is the Fig 1 Title. Prevalence and relative abundance of *D. pigrum* across respiratory health status. This is the Fig 1 legend: (A) Proportion of *D. pigrum* carriers in healthy children, viral and IPD cases. Numbers above bars indicate carrier/total of each group. Differences assessed by Chi-square test. (B) Log-transformed relative abundance of *D. pigrum* in healthy children, viral and IPD cases. (C) Extended comparison including healthy subgroup. Box plots shown median, interquartile range and individual values. Statistical comparisons performed using Kruskal-Wallis test followed by pairwise Wilcoxon tests. P-values are indicated above brackets.

**Table 1:** Epidemiological and clinical characteristics of study population according to presence /absence of *D.pigrum*.

| Variable | Total<br>(n=140) | Presence of<br>*D. pigrum*<br>(n=79) | Absence of<br>*D. pigrum*<br>(n=61) | % of<br>*D. pigrum* |
| --- | --- | --- | --- | --- |
| Age, median months[IQR] | 35.7 [0.1-45.0] | 33.6 [2.0-41.5] | 38.48 [0.1-48.0] |  |
| <2 years | 62 (44.3%) | 36/79(45.6%) | 26/61 (42.6%) | 58.1% |
| >2 years | 78 (55.7%) | 43/79(54.4%) | 35/61(57.3%) | 55.1% |
| Sex |  |  |  |  |
| Male | 73 (52.1%) | 48/79 (60.8%) | 25/61 (41.0%) | 65.8% |
| Female | 67 (47.9%) | 31/79 (39.2%) | 36/61 (59.0%) | 46.3% |
| **Respiratory health status** |  |  |  |  |
| Healthy | 65 (46.4%) | 44/79 (55.7%) | 21/61 (34.4%) | 67.7% |
| Banal Viral cases | 48 (34.3%) | 26/79 (32.9%) | 22/61 (36.1%) | 54.2% |
| IPD cases | 27 (19.3%) | 9/79 (11.4%) | 18/61 (29.5%) | 33.3% |
| **Breastfeeding (n=139)** |  |  |  |  |
| Yes | 110 (79.1%) | 66/79 (83.5%) | 44/60 (73.3%) | 60.0% |
| >6months | 76/110 (69.1%) | 46/66 (69.7%) | 30/44 (68.2%) | 60.5% |
| <=6 months | 34/110 (30.9%) | 20/66 (30.3%) | 14/44(31.8%) | 58.8% |
| No | 29 (20.9%) | 13/79 (16.5%) | 16/60 (26.7%) | 44.8% |
| **Delivery mode (n=133)** |  |  |  |  |
| Vaginal | 92 (69.2%) | 53/76 (69.7%) | 39/57 (68.4%) | 57.6% |
| C-section | 41 (30.8%) | 23/76 (30.3%) | 18/57 (31.6%) | 56.1% |
| **Vaccination status** |  |  |  |  |
| Unvaccinated | 42 (30.0%) | 20/79 (25.3%) | 22/61 (36.1%) | 47.6% |
| Vaccinated | 98 (70.0%) | 59/79 (74.7%) | 39/61 (63.3%) | 60.2% |
| **Kindergarten attendance (n=138)** |  |  |  |  |
| Yes | 65 (47.1%) | 38/78 (48.7%) | 27/60 (45.0%) | 58.5% |
| No | 73 (52.9%) | 40/78 (51.3%) | 33/60 (55.0%) | 54.8% |

Abbreviations: IQR: interquartile range; IPD: Invasive Pneumococcal Disease

Among *D. pigrum* carriers, males predominated (48/79, 60.8%) over females (31/79, 39.2%). Most children were born via vaginal delivery (*D. pigrum* carriers: 69.7%, non-carriers: 68.4%) and were breastfed for more than six months (*D. pigrum* carriers: 69.7%; non-carriers: 68.2%). In relation to the vaccination status, a higher proportion of children with *D. pigrum* were vaccinated (74.7%) than unvaccinated (63.9%). Kindergarten attendance was slightly below 50% in both groups *(Table 1)*.

Regarding clinical features, healthy children represented a proportion of 55.7% in the group with *D. pigrum* and 34.4% in the group without *D. pigrum.* In contrast, IPD represented a proportion of 29.5% among children without *D. pigrum (Table 1).* Among IPD cases, all the children with *D. pigrum* and the majority of the children without *D. pigrum* presented pneumonia (100.0% and 77.8%, respectively) *(Table 2)*.

**Table 2:** Clinical features of IPD cases.

| Variable | Total<br>(n=27) | Presence of<br>*D. pigrum*<br>(n=9) | Absence of<br>*D. pigrum*<br>(n=18) | % of<br>*D. pigrum* |
| --- | --- | --- | --- | --- |
| **Clinical manifestations of IPD cases** |  |  |  |  |
| Bacteraemia | 1 (3.7%) | 0/9 (0%) | 1/18 (5.56%) | 0.0% |
| Pneumonia | 23 (85.2%) | 9/9 (100%) | 14/18 (77.78%) | 39.1% |
| *Non complicated pneumonia* | 7/23 (25.9%) | 4/9 (44.44%) | 3/14 (21.43%) | 57.1% |
| *Complicated pneumonia* | 16/23 (59.3%) | 5/9 (55.56%) | 11/14 (78.57%) | 31.3% |
| Meningitis | 3 (11.1%) | 0/9 (0%) | 3/18 (16.67%) | 0.0% |
| **PICU** |  |  |  |  |
| Yes | 4 (14.8%) | 0/9 (0%) | 4/18 (22.22%) | 0.0% |
| No | 23 (85.2%) | 9/9 (100%) | 14/18 (77.78%) | 39.1% |

Abbreviations: IPD: Invasive Pneumococcal Disease, PICU: Paediatric Intensive Care Unit.

From a microbiological perspective, among participants with the presence of *D. pigrum,* 62.8% were carriers of *S. pneumoniae,* with LRST accounting for 83.7% in the group with *D. pigrum.* Additionally, the children without *D. pigrum* had a 70.5% of pneumococcal carriage *(Table 3).* Respiratory viruses were detected in 74.7% of *D. pigrum* carriers and 86.9% in non-carriers. Rhinovirus/Enterovirus was the most frequent virus found in both groups, followed by Bocavirus (21.4%) in the *D. pigrum*-positive group and Adenovirus (28.3%) in the *D. pigrum*-negative group *(Table 3)*.

**Table 3:**
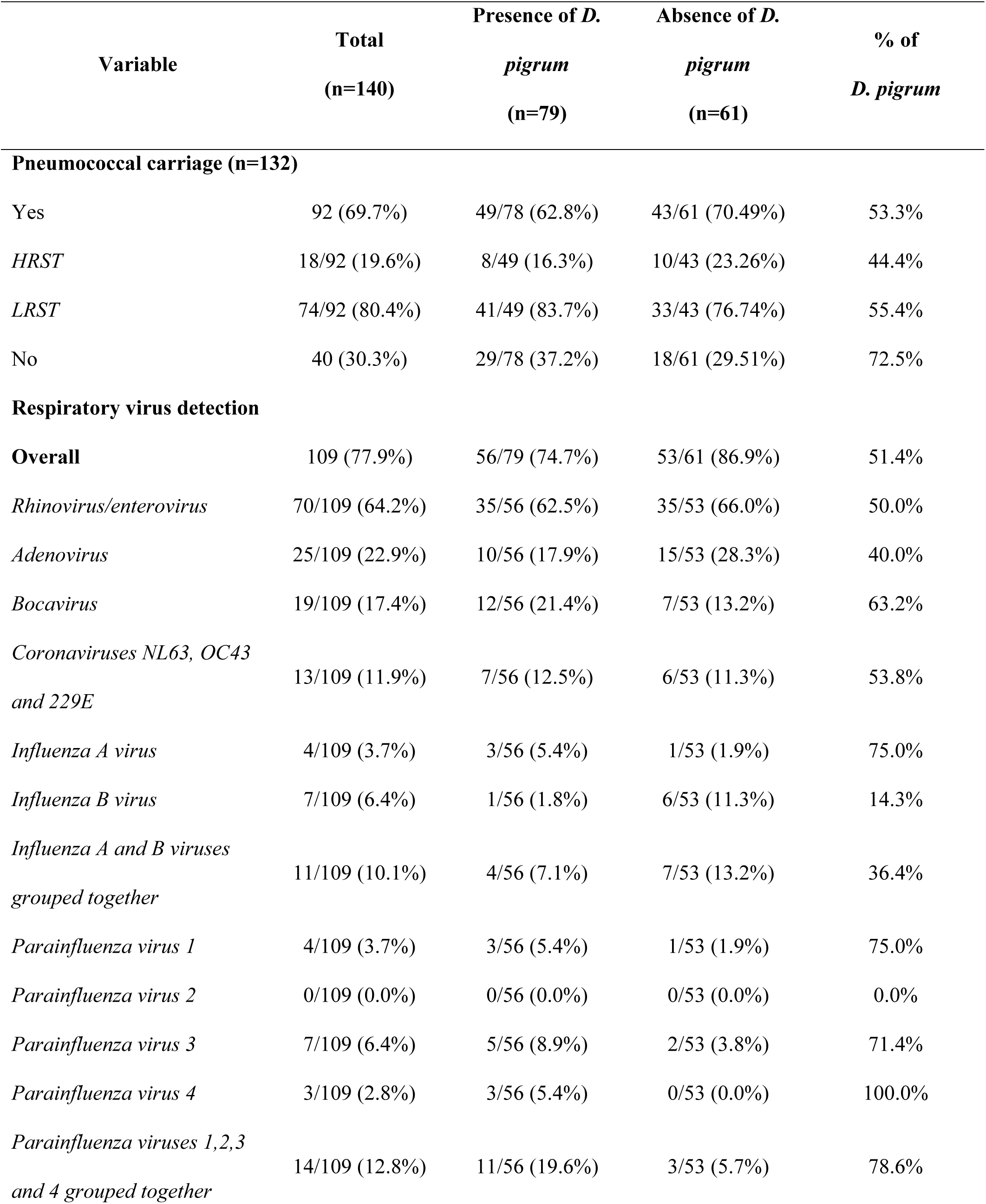
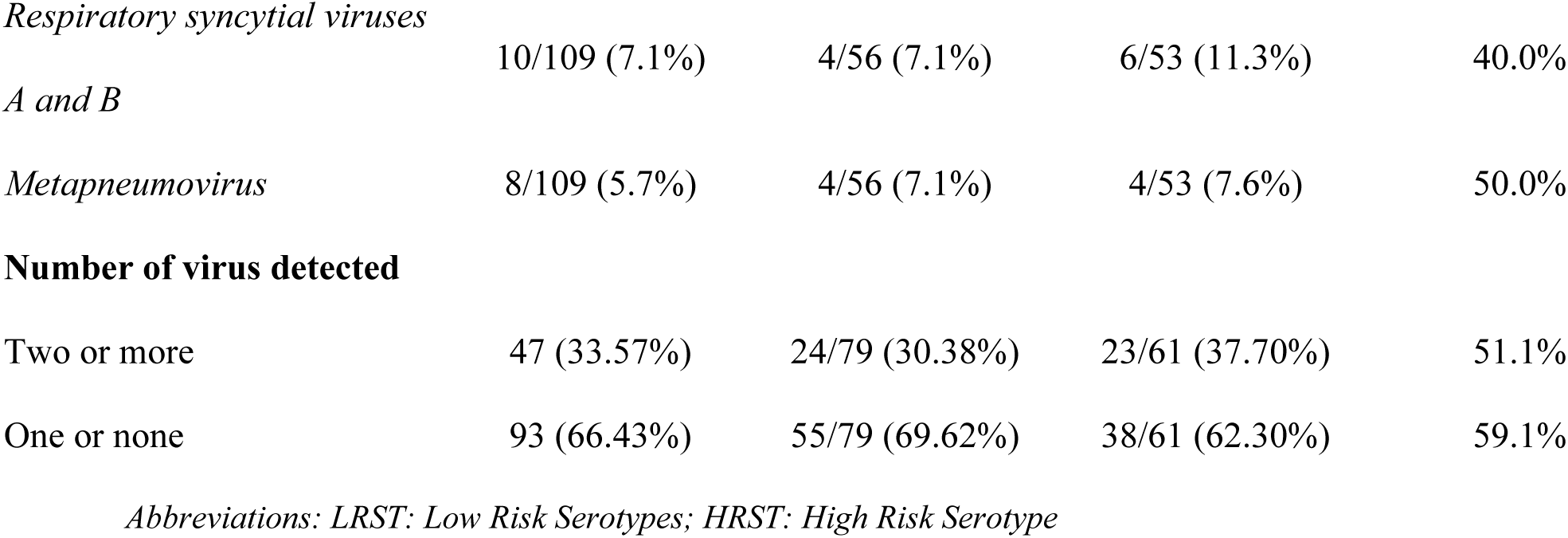
Pneumococcal carriage and respiratory virus detection of study population.

### Association of the presence of D. pigrum with epidemiological, clinical and microbiological variables

Significant differences were observed between males and females in the presence of *D. pigrum*, with males being more likely than to have *D. pigrum* in the nasopharynx (OR: 2.2, 95% CI: 1.12-4.43; p= 0.02). The association remained significant in the multivariate analysis, with an adjusted odds ratio (aOR) of 2.21 (95% CI: 1.07–4.6; p= 0.03). Children who were not breastfed had a lower probability of having *D. pigrum* compared to those who were breastfed, but the difference between these proportions was not statistically significant (univariate, p= 0.15 and multivariate p= 0.06). No significant associations were found between *D. pigrum* colonisation and age group, type of delivery, vaccination status and kindergarten attendance.

The univariate analysis revealed a significant association of the respiratory health status and *D. pigrum* colonisation, with healthy children more likely to carry *D. pigrum* compared with those with IPD (OR, 4.1; 95% CI: 1.6–11.5; p= 0.003). This association remained significant in the multivariate analysis, with an aOR of 3.7 (95% CI: 1.1–12.6; p= 0.028). Both univariate and multivariate analyses showed no significant differences were observed between healthy children and viral cases (univariate, p= 0.15 and multivariate p= 0.86) or between viral and IPD cases (univariate analysis: p= 0.09; multivariate analysis: p= 0.10) *(Table 4)*.

**Table 4:** Factors related with the presence of D. pigrum.

| Predictor variables | Univariate analysis |  |  | Multivariate analysis |  |  |
| --- | --- | --- | --- | --- | --- | --- |
|  | OR | 95% CI | p-value | aOR | 95% CI | p-value |
| Age group, <2 years vs. >2 years | 1.125 | 0.572-2.224 | 0.732 |  |  |  |
| Sex, male vs. female | 2.212 | 1.122-4.431 | 0.022 | 2.21 | 1.068-4.566 | 0.033 |
| Respiratory health status, healthy vs. IPD | 4.090 | 1.598-11.154 | 0.003 | 3.756 | 1.121-12.59 | 0.028 |
| Respiratory health status healthy vs. viral | 1.762 | 0.813-3.854 | 0.151 | 1.266 | 0.447- 3.58 | 0.856 |
| Respiratory health status, viral vs. IPD | 2.322 | 0.878-6.483 | 0.09 | 2.968 | 0.847-10.309 | 0.103 |
| Breastfeeding, yes vs no | 1.834 | 0.799-4.280 | 0.152 | 2.33 | 0.950-5.680 | 0.063 |
| Delivery mode, vaginal vs. c-section | 1.064 | 0.500-2.245 | 0.871 |  |  |  |
| Kindergarten attendance, no vs. yes | 0.863 | 0.436-1.699 | 0.669 |  |  |  |
| Vaccination status, vaccinated<br>vs. not vaccinated | 1.656 | 0.797-3.471 | 0.177 |  |  |  |
| Pneumococcal carriage,<br>negative vs. positive | 1.407 | 0.688-2.929 | 0.351 |  |  |  |
| Respiratory virus detection,<br>negative vs. positive | 2.673 | 1.130-6.934 | 0.024 | 2.67 | 0.962-7.44 | 0.059 |
| Rhinovirus/enterovirus,<br>negative vs. positive | 1.684 | 0.858-3.339 | 0.130 |  |  |  |
| Adenovirus, negative vs.<br>positive | 2.226 | 0.923-5.588 | 0.075 |  |  |  |
| Bocavirus, negative vs. positive | 0.759 | 0.262-2.049 | 0.591 |  |  |  |
| Coronaviruses NL63, OC43<br>and 229E | 1.125 | 0.336-3.650 | 0.844 |  |  |  |
| Influenza A and B viruses<br>grouped together | 2.423 | 0.678-10.022 | 0.207 |  |  |  |
| Parainfluenza viruses 1,2,3 and<br>4 grouped together | 0.333 | 0.069-1.148 | 0.084 |  |  |  |
| Respiratory syncytial viruses A<br>and B | 2.048 | 0.542-8.691 | 0.328 |  |  |  |
| Metapneumovirus | 1.313 | 0.284-6.052 | 0.728 |  |  |  |

Abbreviations: OR: Odds Ratio; CI: Confidence Interval; aOR: adjusted Odds Ratio; IPD: Invasive Pneumococcal Disease
Note: In all contrasts, the first category represents the comparison group (numerator) and the second category is the reference group (denominator). Odds ratios (OR) indicate the odds of *D. pigrum* carriage in the first group relative to the second

Absence of respiratory viruses was associated with a higher proportion of *D. pigrum’s* presence in the univariate analysis (OR: 2.7; 95% CI: 1.1-6.9; p =0.02), but not in the multivariate analysis (p=0.06). The distribution of individual respiratory viruses did not differ significantly across *D. pigrum* carriers and non-carriers. No significant differences were found in relation to the presence of *D. pigrum* according to *S. pneumoniae* carrier status *(Table 4)*.

### Differential abundance of *D. pigrum* across respiratory health status groups

Relative abundance of *D. pigrum* was evaluated across all participants as a complementary analysis to assess the effect of the abundance beyond presence/absence. This analysis showed significantly higher abundance of *D. pigrum* among healthy children compared to IPD cases (p= 0.001) (*Fig. 1B*). Viral cases exhibited an intermediate level of *D. pigrum* abundance, with significant differences compared to healthy children (p= 0.029) but not when compared to IPD cases (p= 0.06). Further subclassification of healthy children revealed significant differences in *D. pigrum* abundance between subgroups (p= 0.003), with the healthy virus-negative group showing the highest abundance (*Fig. 1C*). This group also showed significantly higher *D. pigrum* abundance than viral cases (p= 0.001) and IPD cases (p= 0.0002). In contrast, no significant differences were observed between healthy virus-positive carriers and either viral or IPD cases.

## DISCUSSION

This study examined epidemiological, clinical, and microbiological variables associated with *Dolosigranulum pigrum* presence in the paediatric nasopharynx during the pre-pandemic period. Given the emerging evidence of the potential protective role in respiratory health of *D. pigrum* [8–14], the study included different study groups according to different respiratory health statuses to provide a comprehensive analysis.

Results demonstrated a significant association between *D. pigrum* presence and male sex, with males more likely to have nasopharyngeal *D. pigrum* colonisation. This finding sheds more evidence of sex-based differences in bacterial composition [38,39]. Previous research has suggested that the human microbiome is influenced by hormonal, genetic, and behavioural factors [40,41], which vary between sexes. These differences could impact the composition of the respiratory microbiota, potentially allowing *D. pigrum* to play a compensatory role in protecting against pathogens in males, who are known to have a higher risk of lower respiratory tract infections (LTRIs) [40,42]. Although this hypothesis has not been deeply investigated, it raises the possibility that the presence of *D. pigrum* in males could represent an evolutionary ecological adaptation to balance sex disparities in disease susceptibility. Furthermore, the lower prevalence of *S. pneumoniae* carriers among males reported in a retrospective cohort study [43] could contribute to the higher prevalence of *D. pigrum* in this study group, given their antagonistic relationship [14,21]. Despite these insights, research on sex-related microbiota differences remains in its early stages, with limited evidence currently available. The present study contributes to this emerging area of research by providing additional evidence of sex-related microbiome variations.

Moreover, the findings revealed a significant association with the presence of *D. pigrum* and respiratory health status. Specifically, healthy children were more likely to have *D. pigrum* compared to IPD cases. These findings are aligned with the hypothesis that *D. pigrum* may play a protective role against IPD, a condition caused by the invasion of *S. pneumoniae* into sterile territories [2,3]. As previously reported pneumococcal colonisation is considered a major risk for IPD [2], consequently in this study *S. pneumoniae* carrier status was assessed in all participants. Although a lower proportion of pneumococcal carriers was noted among children with *D. pigrum*, the association did not reach statistical significance. This suggests that the protective role of *D. pigrum* may extend beyond inhibiting pneumococcal colonisation, as previously reported [14,21]. It is plausible that *D. pigrum* employs additional mechanisms that impede the transition of *S. pneumoniae* from a colonising to an invasive state, therefore playing a role in preventing IPD. These mechanisms could include interactions with other microbiome components [14,21], modulation of the host immune response [22] or the production of metabolites that interfere with the virulence of *S. pneumoniae* [44–46]. Furthermore, the analysis of *D. pigrum* relative abundance provided additional evidence to these findings supporting this protective role. Healthy children not only had higher prevalence of *D. pigrum* but also showed greater abundance compared to IPD cases. This dose-dependent pattern suggests that the colonisation density of *D. pigrum* might be related with its protective role, with higher bacterial loads potentially conferring stronger resistance against progression to invasive disease.

In contrast, regarding respiratory health status, no significant association was observed between *D. pigrum* presence and viral infection. In addition, the association between the presence of *D. pigrum* and PCR-confirmed detection of respiratory viruses was evaluated. This analysis demonstrated that children with *D. pigrum* were less likely to have respiratory viruses. Although this association reached statistical significance in the univariate analysis, it was not found in the multivariate analysis, remaining as a non-significant trend. This attenuation effect in the multivariate model may reflect the influence of confounding variables, suggesting that the univariate association could be explained by other covariates rather than a direct role of *D. pigrum* presence. However, across all study groups, respiratory virus infections were banal or asymptomatic. This result aligns with previous reports suggesting that severe respiratory infections are typically associated with the absence of *D. pigrum* [25]. However, the relative abundance analysis showed that the presence of respiratory virus significantly affects *D. pigrum* colonisation, resulting in lower abundance. This finding suggests that respiratory viruses influence the colonisation density of *D. pigrum* in the nasopharynx. Nevertheless, all these findings highlight the compelling necessity of future studies to clarify the potential role of *D. pigrum* in modulating the respiratory microbiome and its implications for paediatric respiratory health.

The present study has some limitations. Firstly, the sample size of the IPD case study group was smaller compared to the other study groups. This limitation derives from the selection criterion of including only cases with less than 24 hours of antibiotic exposure. Many potential IPD cases were excluded because they had received antibiotic treatment for longer periods before sample collection, which would have compromised the reliability of the results. Given that *D. pigrum* is a gram-positive coccus highly sensitive to antibiotics such as beta-lactams[16], prolonged antibiotic exposure could have reduced its detectable presence, leading to false-negative results. Therefore, ensuring this criterion was crucial to maintain the reliability and robustness of the findings.

However, the reduced sample size of our study highlights the need for future studies with larger cohorts to confirm and expand these observations. Secondly, the relative abundance of *D. pigrum* was calculated as the proportion of reads specifically assigned to this specific taxon relative to the total number of reads obtained by sequencing the V3-V4 region of the 16S rRNA gene. While this approach is common in microbiota studies, it presents specific challenges, such as the low taxonomic resolution of the 16S rRNA gene amplicon, which may limit the accurate identification of *D. pigrum* at the species level. Therefore, future studies could complement this approach with 3^rd^ generation full-length 16S sequencing [47], WGS metagenomics data or quantitative techniques (such as qPCR) to validate and refine the determination of bacterial presence. Thirdly, although previous studies have demonstrated a synergistic effect between *D. pigrum* and *C. pseudodiphtheriticum* in the reduction of pneumococcal growth [14], the present study focused exclusively on *D. pigrum*. Future studies should explore these bacteria interactions to better understand the complex ecological dynamics that may contribute to respiratory health.

In summary, the present study provides further evidence supporting the protective role of *D. pigrum* against IPD, highlighting its potential as a key modulator of paediatric respiratory health. These findings reinforce the importance of future studies to elucidate the underlying mechanisms and to evaluate the potential for leveraging *D. pigrum* in preventive or therapeutic strategies against severe respiratory infections.

## FUNDING STATEMENT

The authors acknowledge the financial assistance extended by the projects FIS PI13/01884, PI16/00174, PI19/00104 and PI23/00049, to PFIS fellowship FI 24/00206.

This support was crucial for the attainment of results presented in the present research.

## ETHICAL APPROVAL STATEMENT

This is an ancillary study of data from a previously published study [20] .The original research was approved by the Ethics Committee of Hospital Sant Joan de Déu (PIC 70-15 and PIC 137-16) and conducted in compliance with the Helsinki Declaration and Spanish regulations on data protection (Organic Laws 15/1999 and 14/2007). Written informed consent was obtained from all parents/legal guardians in the original study. Data used in the present analysis were anonymized.

## AUTHOR CONTRIBUTIONS

Cisneros M: Formal analysis/Investigation/Visualization/ Writing-original draft

Henares D: Data curation/Methodology/ Writing-review and editing

Lluansi A: Methodology /Writing-review and editing

Brotons P: Methodology/Writing-review and editing

Launes C: Writing-review and editing

Penela-Sanchez D: Writing-review and editing

Gonzalez-Comino G: Writing-review and editing

Perez-Argüello A: Data curation/ Writing-review and editing

de Sevilla MF: Writing-review and editing

Mira A: Methodology/Writing-original draft

Blanco-Fuertes M: Conceptualization /Formal analysis/Supervision/ Writing-original draft

Muñoz-Almagro C: Conceptualization/ Funding acquisition/Project administration/Supervision /Writing-original draft

## Data Availability

All data used in this study are publicly available. The dataset can be found in the supplementary materials of the original publication (Appendix S1) and is available at Figshare: https://doi.org/10.6084/m9.figshare.13280435.v3.

https://doi.org/10.6084/m9.figshare.13280435.v3.

## Notes

### Competing Interest Statement

I have read the journal's policy and the authors of this manuscript have the following competing interests: Carmen Munoz-Almagro reports travel grants from Pfizer, MSD, and personal fees as speaker from Sanofi-Pasteur and MSD. The rest of authors declare no conflicts of interest.

### Funding Statement

PI13/01884
PI16/00174
PI19/00104
PI23/00049
FI24/00206

### Author Declarations

Ethics Committee of Hospital Sant Joan de DEu gave ethical approval for the original study from which these data were obtained. The present study is an ancillary analysis of that dataset.

